# A computationally efficient approach to segmentation of the aorta and coronary arteries using deep learning

**DOI:** 10.1101/2021.02.18.21252005

**Authors:** Wing Keung Cheung, Robert Bell, Arjun Nair, Leon Menezies, Riyaz Patel, Simon Wan, Kacy Chou, Jiahang Chen, Ryo Torii, Rhodri H. Davies, James C. Moon, Daniel C. Alexander, Joseph Jacob

## Abstract

A fully automatic two-dimensional Unet model is proposed to segment aorta and coronary arteries in computed tomography images. Two models are trained to segment two regions of interest, (1) the aorta and the coronary arteries or (2) the coronary arteries alone. Our method achieves 91.20% and 88.80% dice similarity coefficient accuracy on regions of interest 1 and 2 respectively. Compared with a semi-automatic segmentation method, our model performs better when segmenting the coronary arteries alone. The performance of the proposed method is comparable to existing published two-dimensional or three-dimensional deep learning models. Furthermore, the algorithmic and graphical processing unit memory efficiencies are maintained such that the model can be deployed within hospital computer networks where graphical processing units are typically not available.

## Introduction

### Background

Coronary artery disease (CAD) is one of the leading causes of death in the UK [1] and worldwide. The lumen of the coronary arteries can narrow as a result of build-up of atheromatous plaque within the artery wall. Reductions in local blood flow as a result of vessel lumen narrowing can starve heart muscle of oxygen. Vulnerable plaque can rupture, occlude the vessel lumen and result in a cardiac muscle ischaemia/death, which manifests clinically as a heart attack. Early detection of the presence of atheromatous plaque and vessel stenosis [2] could allow early medical intervention and potentially reduce the risk of heart attack. Currently, several non-invasive imaging modalities are available to clinicians for visualising the anatomy of the coronary arteries as well as delineating the severity of vessel stenosis. These imaging modalities are Stress Echocardiography [3], Cardiac magnetic resonance imaging (MRI) [4] and Computed Tomography Coronary Angiography (CTCA) [5]. CTCA is the quickest of these methods and offers high sensitivity and specificity for detection and exclusion of significant coronary stenosis [6]. As a result CTCA is the preferred first-line option for the assessment of stable cardiac disease in the National Institute for Health and Care Excellences guidelines for the UK [7]. CTCA has the ability to identify calcified, non-calcified coronary plaque and mixed-attenuation plaques which can help clinicians characterise plaques and formulate management strategies.

To assess the severity of CAD, one approach involves visual estimation of stenosis severity on CTCA scans. This requires the geometrical information of the coronary arteries to be provided in order to accurately assess the severity of the stenosis. This approach however is subjective and time consuming. Accurate interpretation and diagnosis of CAD is heavily reliant on the experience and expertise of individual clinicians [8, 9]. The diagnostic outcomes can differ between newly trained clinicians compared to experienced specialists.

An alternative approach for assessing CAD severity involves performing computational fluid dynamics (CFD) on the target arteries [10]. It first requires identification of accurate geometries of the aorta and coronary arteries. A set of partial differential equations of blood flow are then solved numerically given the boundary conditions and the geometries. Once the blood flow is estimated, the useful clinical predictor, Fractional Flow Reserve (FFR) [11-13] can be derived. This approach provides an objective estimation of stenosis and does not require additional imaging, which makes it particularly attractive to clinicians. However, implementing CFD models is very time consuming and computationally demanding. It is usually deployed on high performance computer clusters (HPC) [14] and takes several hours per patient to produce the results. Importantly, the CFD simulation has to be performed off-site where HPC services are available. This approach is not practical for real-time patient management.

Deriving accurate geometrical information of the aorta and coronary arteries is important for the above approaches. It is achieved by delineating the outline of the vessels, termed segmentation. Blood vessel segmentation can be performed manually, semi-automatically or automatically [15-20]. Manual segmentation is subjective and time consuming, requiring pixel-by-pixel labelling of individual vessels. Semi-automatic and automatic segmentation methods are objective and quicker, though they can require manual correction for under- or over-segmented vessels. There remains an unmet need to develop fast, objective and accurate automated computer-derived coronary artery segmentation algorithms that can be deployed in a hospital setting to assist clinicians diagnose CAD. This is especially relevant in Accident and Emergency (A+E) departments where CTCA reviews are often delayed due to a lack of available specialists to read the CTCAs [21]. This delay in turn slows patient management in A+E, increases resource utilisation and results in excess costs to the health service.

A fully automatic detection and classification system for CAD using computer-based deep learning algorithms is a way to achieve the above goal. The first stage of the process initially requires segmentation (identifying the outlines) of the aorta and coronary arteries on CTCA images. The second stage involves classification of disease severity performed on segmented CTCA images. The segmentation task has be considered in two ways in the literature. The first involves segmentation of both the aorta and the coronary arteries. For example, Gu et al [22] proposed a 3D deep learning model to perform this task. The other strategy is to segment just the coronary arteries alone. For example, Huang et al [23] suggested a 3D deep learning method with centreline to segment the coronary arteries. In general, the performance of aorta and coronary artery segmentation is better than segmentation of the coronary arteries alone. A more detailed summary regarding existing deep learning-based segmentations is discussed in the following paragraph.

The works related to artery segmentation on CTCA images have been discussed in two review papers [20, 24]. The focus of the current study relates to deep learning methods of coronary artery segmentation. Therefore, we briefly summarise the work published so far. Several deep learning techniques have been proposed to segment the aorta and/or coronary arteries. Moeskops et al [25] investigated a single convolutional neural network trained to segment coronary arteries in cardiac CTA images. The training dice similarity coefficient (DSC) accuracy was around 65%. A 3D-convolutional neural network was presented by Merkow et al [26], which demonstrated that processing the volumetric data in 3D could improve the segmentation performance compared to 2D processing. However, the performance of the coronary artery segmentation model was not reported. Kjerland et al [27] adopted a 3D DeepMedic network to segment both the aorta and coronary arteries. The reported DSC accuracy was between 75%-78%. Huang et al [23] examined a 3D Unet with/without a centerline to segment the coronary artery. The DSC accuracy was between 71%-78%.

Recently, a 3D multi-channel Unet has been proposed by Chen et al [28], which had a DSC accuracy of 80% for coronary artery segmentation. Shen et al [29] proposed a 3D fully convolutional network with attention gates to segment both the aorta and coronary artery. The boundary of the segmented artery was smoothed by a level set function. The average DSC accuracy was about 90%. Lee et al [30] introduced a template transformer network where a shape template is deformed to match the underlying structure of interest through an end-to-end trained spatial transformer network for coronary artery segmentation. The DSC accuracy is between 76%-78%. Wolterink et al [31] proposed using graph convolutional networks to predict the spatial location of vertices in a tubular surface mesh that segments the coronary artery lumen. The average DSC is 74%. Mirunalini et al [32] proposed a two-stage approach to segment the coronary artery. The first stage adopted a 2D Recurrent Convolutional Neural Network to detect the artery in the slice, then a 2D residual Unet was used to segment the coronary artery. The intersection over union (IoU) was reported, which was 84%. Lei et al [33] developed a 3D Attention Fully Convolutional Network model to automatically segment the aorta and coronary artery for CCTA. The mean DSC is 83%. Gu et al [22] recently published a 3D global feature embedded network with active contour loss to segment the aorta and coronary artery. The reported average DSC is 91.43%.

In this study, we propose a modified U-Net [34] model and evaluate its performance for the automated segmentation of the aorta and coronary arteries on CTCA images. We then retrain our model to segment the coronary arteries alone and demonstrate improved performance.

The main contributions of this study are:

- The first study to propose a modified 2D Unet directly to segment the aorta and/or coronary arteries in CTCA scans
- This fully automated technique is simple, fast and efficient, and produces results that can assist real-time clinical decisions
- It is practically feasible to be implemented in clinical systems where available computational resources are limited
- The performance of this model is similar to other existing deep learning techniques (3D global feature embedded network + Active contour loss) (with aorta) / (2D RCNN + 2D Unet) (without aorta)
- Importantly, our technique works well when the coronary arteries alone are segmented (accuracy ∼89%).

## Method

### Clinical data

CTCA scans were performed on 69 subjects and patients with chest pain. The scans were acquired at University College Hospital London and Barts Health NHS Trust using different CT scanners and acquisition protocols. An example of a CTCA scan is shown in Figure 1.

**Figure 1:**
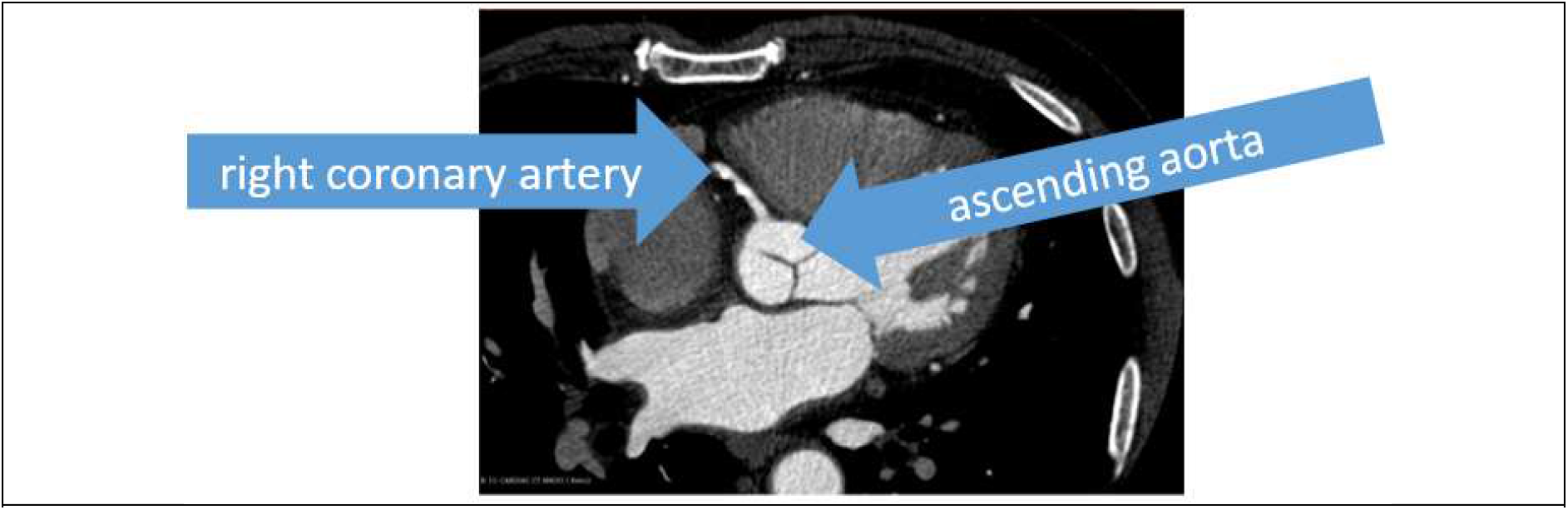
A CTCA scan of a patient

### Data pre-processing

The original Digital Imaging and Communications in Medicine (DICOM) data were pre-processed using ImageJ [35]. The image size was 512 x 512 pixels. The pixel intensity was normalised by using linear histogram stretch and then rescaled to between 0 to 255. Final images were converted to 8-bit Portable Network Graphics (PNG) for training, validation and testing.

### Semi-automatic segmentation

Initial annotation was performed using Simpleware-ScanIP (Version 2018.12; Synopsys, Inc., Mountain View, USA). The segmentation procedure consisted of thresholding, background flood-fill and split algorithms. Firstly, the thresholding was applied such that only regions containing contrast were considered. Secondly, a seed point was placed within the aorta, and the background flood-fill algorithm was able to segment the coronary arteries and cardiac chambers which were connected to the aorta. Lastly, the split algorithm was performed such that the aorta and the coronary arteries were separated from the cardiac chambers. It should be noted that the split operation may be repeated such that all connected chambers are separated. The workflow of these procedures is displayed in Figure 2.

**Figure 2:**
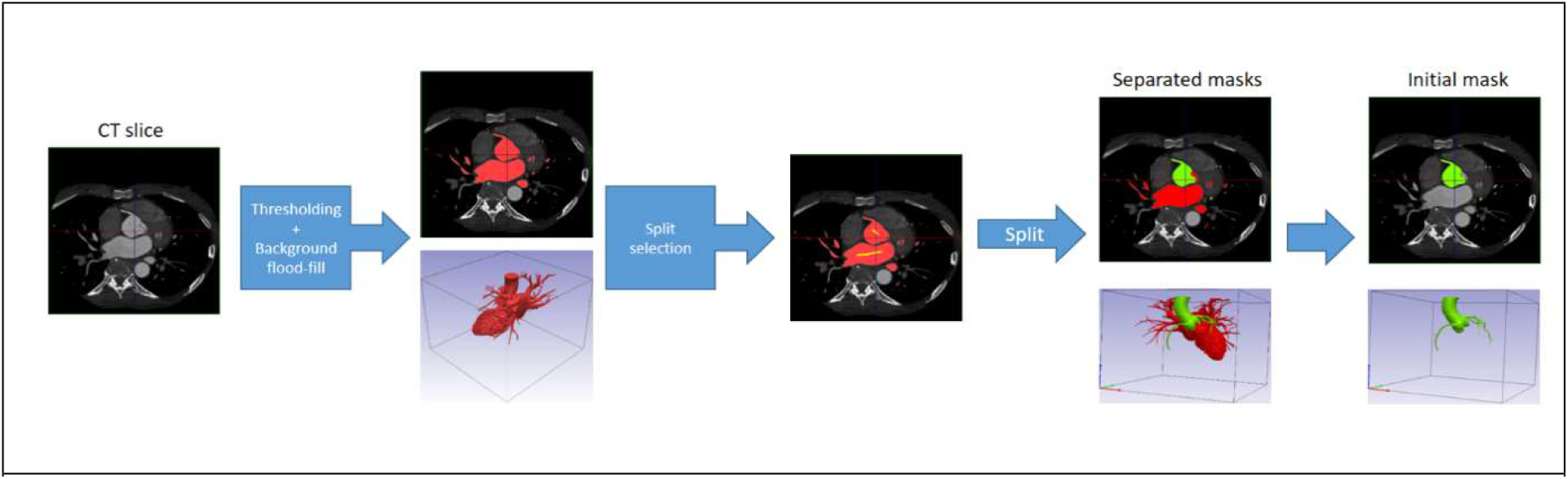
The workflow of initial annotation by using Simpleware-ScanIP

The initial mask contained the ascending aorta (AA), right coronary artery (RCA), left circumflex artery (LCX) and left coronary artery (LCA). The mask was then fine-tuned manually using 3D Slicer [36]. To acquire the mask containing just the coronary arteries, the AA was removed to leave the RCA, LCX and LCA only. An example of initial and final masks is shown in Figure 3.

**Figure 3:**
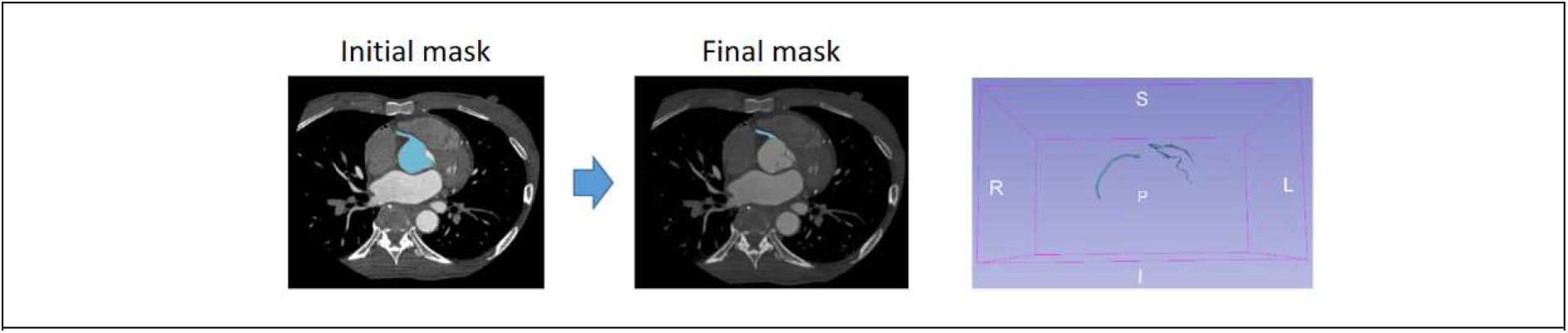
The final coronary artery mask fine-tuned using 3D Slicer

### Manual segmentation

The manual segmentation was implemented in Slicer 3D. The annotator highlighted the vessel by identifying the contrast within the CTCA image given an initial mask. The segmented masks were used as the optimum manual (ground-truth) labels.

### Segmentation methods

#### (1) Aorta and coronary artery segmentation

The regions of interest that the current work focusses on are the aorta and coronary arteries. There are two scenarios that these masks are used. (1) The combined mask of the aorta and coronary arteries is useful for blood flow estimation by using computational fluid dynamics. Aortic segmentations can produce continuous 3D measurements of aortic size and shape which are objective and allow detailed longitudinal comparisons of subtle changes in aortic morphology. (2) The mask of coronary arteries alone is useful for cardiologists to assess the degree of stenosis in areas where CAD has developed. Therefore, the proposed method was evaluated on these two segmented masks.

#### (2) Fully automatic segmentation

##### (A) Our proposed model

Our model is based on the 2D Unet [34]. A Unet is a deep convolutional neural network consisting of down-sample and up-sample paths. The first component of the network extracts spatial features and contexts, while the second component localizes the features by using transposed convolutions. A sigmoid function is used for the final background/foreground classification. We have modified the Unet model in two ways: (1) a batch normalization layer is added to the convolution block; (2) a dropout layer is added before each convolution block. This additional implementation improves the stability and performance of the proposed model. The details of our proposed model is shown in Figure 4.

**Figure 4:**
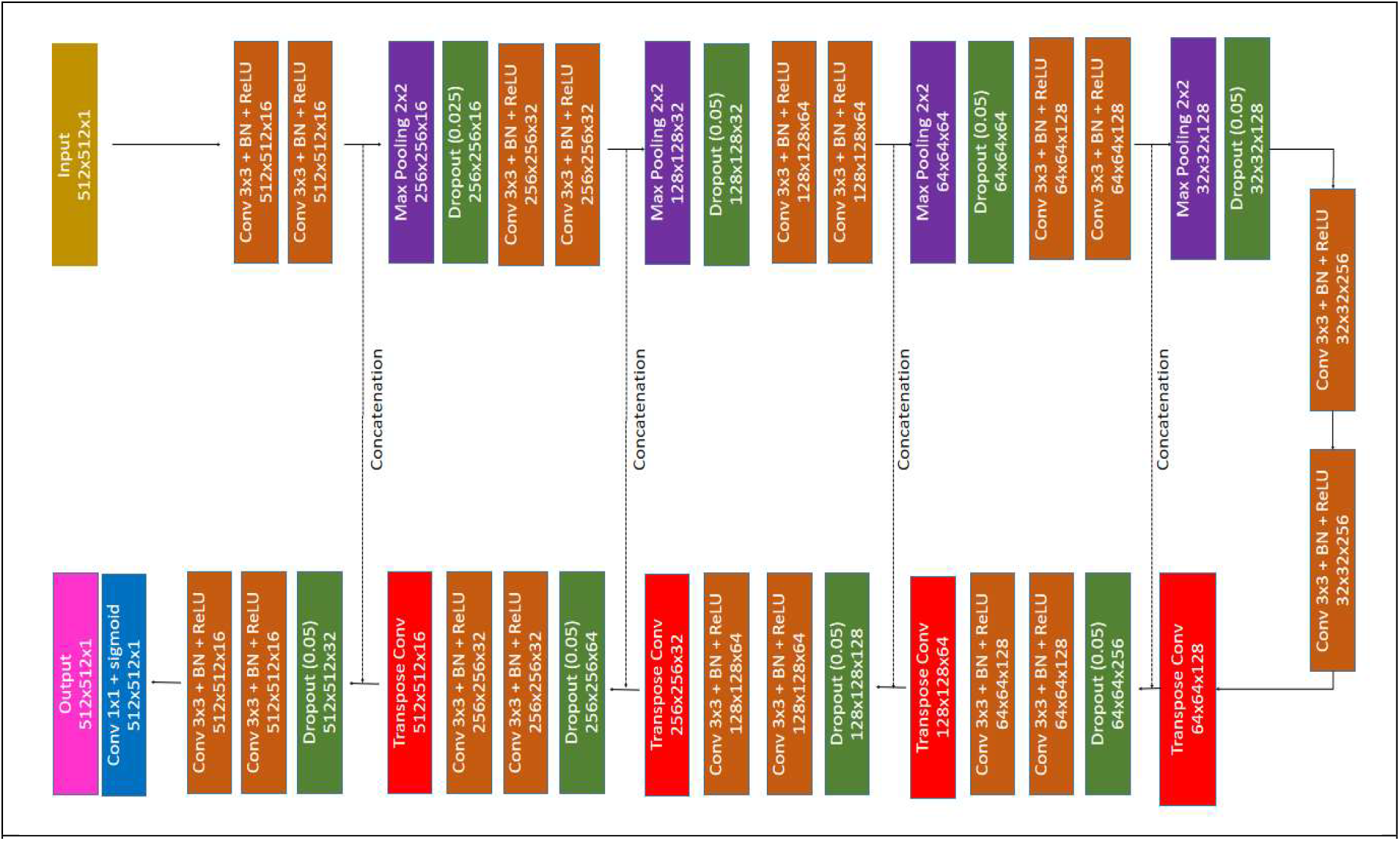
The network architecture of our proposed method

##### (B) Model training

55 datasets (n=13 with no coronary disease, n=42 with coronary disease) were used for training (80%) and validation (20%) The test data set contained 14 datasets (n=5 with no coronary disease, n=9 with coronary disease). There were 11677 slices in the training dataset and 2920 slices in the validation dataset. Slice by slice training was adopted. Two models were trained by using the following optimum manual labels: (1) Aorta and coronary arteries (2) coronary arteries only.

##### (C) Training implementation

The proposed models were implemented in Tensorflow (v 2.1.0) and Keras (2.3.1) on Linux (Rocks 7). They were executed on a cluster (Intel Xeon Gold 5118, 2.3GHz) with a Tesla V100-PCIE-32GB GPU. The Adam algorithm was used to optimise the proposed models. The learning rate was initially set to 1e-5. 200 epochs were set for model training. Early stopping was executed when the loss was not reduced across 10 consecutive epochs.

##### (D) Loss function and performance evaluation

The combined binary cross entropy (BCE) and dice similarity coefficient (DSC) with equal weight were used as the loss function for deep learning. The segmentation performance was measured by using DSC and IoU metrics which are commonly used to measure the similarity between two segmentations.

##### (E) The segmentation prediction implementation

The prediction was performed by using the trained models above. It was implemented on Tensorflow (v 2.1.0) and Keras (v 2.3.1) on Windows 10 and executed on a machine (Intel i9-9960X, 3.1GHz) with a Nvidia Geforce RTX 2700 GPU. The time required for the prediction was also recorded on a per subject/patient basis.

### Segmentation performance and time evaluation

The accuracy of the segmentation performance of our proposed method was compared with published accuracies of existing 2D and 3D deep learning models. For the test dataset, the time required for segmentation and the segmentation performance for our method was compared to semi-automatic segmentation methods. The performance of the segmentation was evaluated by using the DSC and IoU metrics. The Mann-Whitney U Test was performed to evaluate whether there was any difference in segmentation time between automatic and semi-automatic approaches. The analyses were implemented on SPSS (IBM SPSS Statistics for Windows, version 25, IBM, Armonk, NY, USA).

### Experiments and results

#### Learning curve

The learning curves of our model for two scenarios are shown in Figure 5. No overfitting was found in the training for both scenarios. Some fluctuations of the loss function were found at an early stage of training, though the training became stable later on. This potentially reflects the fact that the training was performed in mini-batches. The training that used the aorta and coronary arteries as ground-truth labels took 125 epochs, while the training using coronary arteries alone as the ground-truth label took only 51 epochs. This indicates that the aorta and coronary arteries have distinct features that took longer to learn in the first scenario.

**Figure 5:**
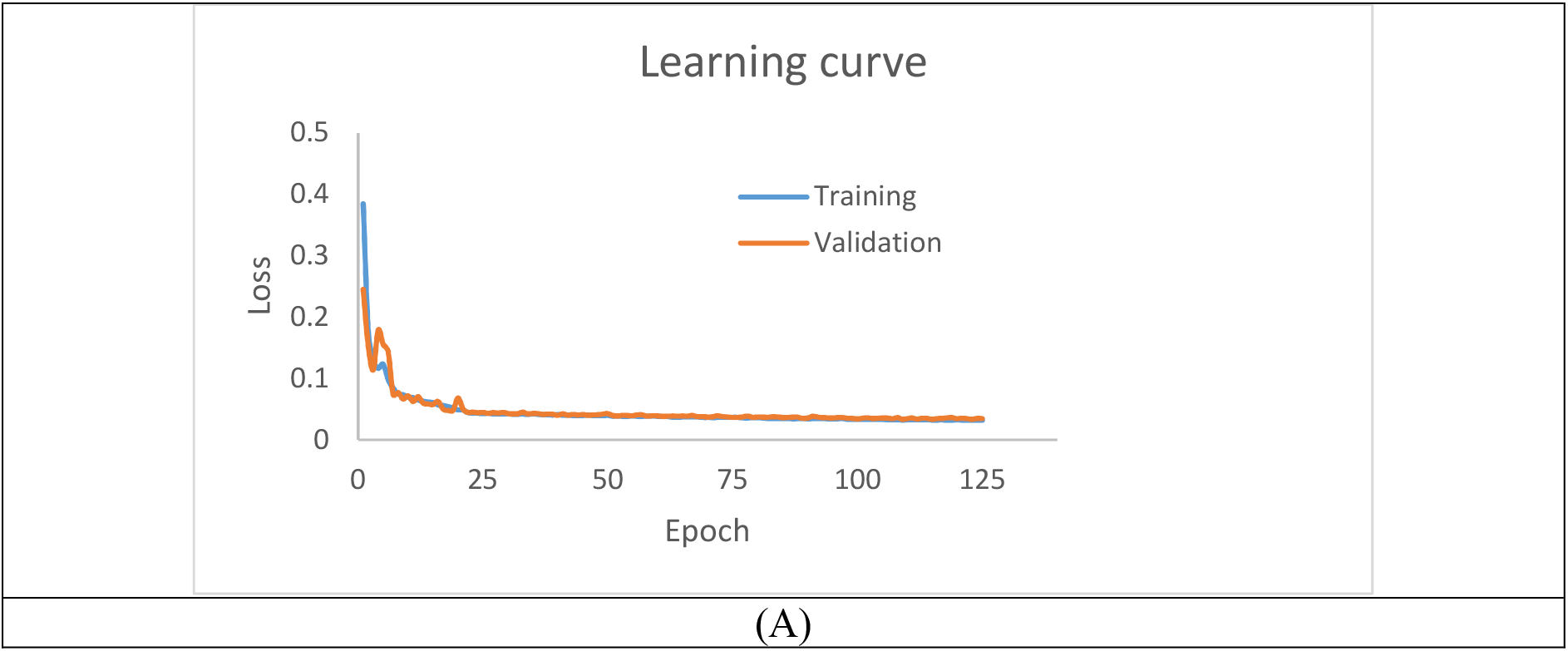
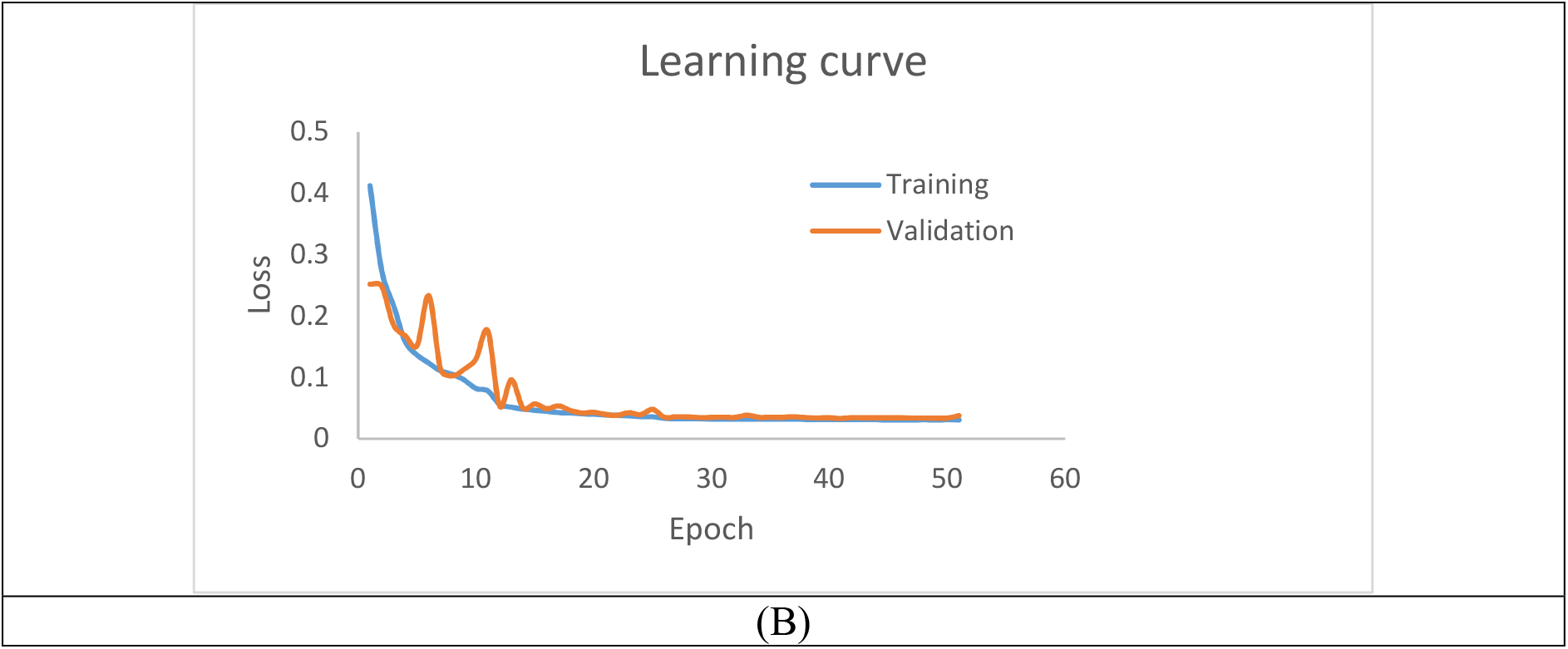
Learning curves: trained with (A) aorta and coronary arteries (B) coronary arteries only

#### Segmentation performance

Table 1 shows the segmentation performance when the aorta and coronary arteries were segmented. The accuracy of our method and Simpleware-ScanIP are 91.20% and 99.40% respectively. The semi-automatic approach performed better than our method when both the aorta and coronary arteries were present in the mask.

**Table 1:** Segmentation performance – mask contains aorta and coronary arteries

|  |  |  |
| --- | --- | --- |
| With aorta | DSC (average) | IoU (average) |
| Simpleware-ScanIP | 99.40% | 98.81% |
| Our method | 91.20% | 83.82% |

The performance of segmentation of the coronary arteries alone is shown in Table 2. The accuracy of our method and Simpleware-ScanIP were 88.80% and 73.22% respectively. Our method performs better than the semi-automatic approach when just the coronary arteries are present in the mask.

**Table 2:** Segmentation performance – mask contains coronary arteries only

|  |  |  |
| --- | --- | --- |
| Without aorta | DSC ( average) | IoU ( average) |
| Simpleware-ScanIP | 73.22% | 57.75% |
| Our method | 88.80% | 79.85% |

The results demonstrate that a semi-automatic approach is good at segmenting the aorta. The semi-automatic approach was limited in its ability to segment the coronary arteries, but as the aorta occupied most of the volume of the mask, the overall segmentation accuracy remained high. Our method performed well when attempting to segment the coronary arteries alone. This suggests that our model has the ability to utilise other features (i.e. shape) to recognise the coronary arteries, while the semi-automatic approach relies solely on pixel density. If the contrast within the coronary artery is not bright enough, the semi-automatic approach will miss some segments of the coronary artery (See Figure 6).

**Figure 6:**
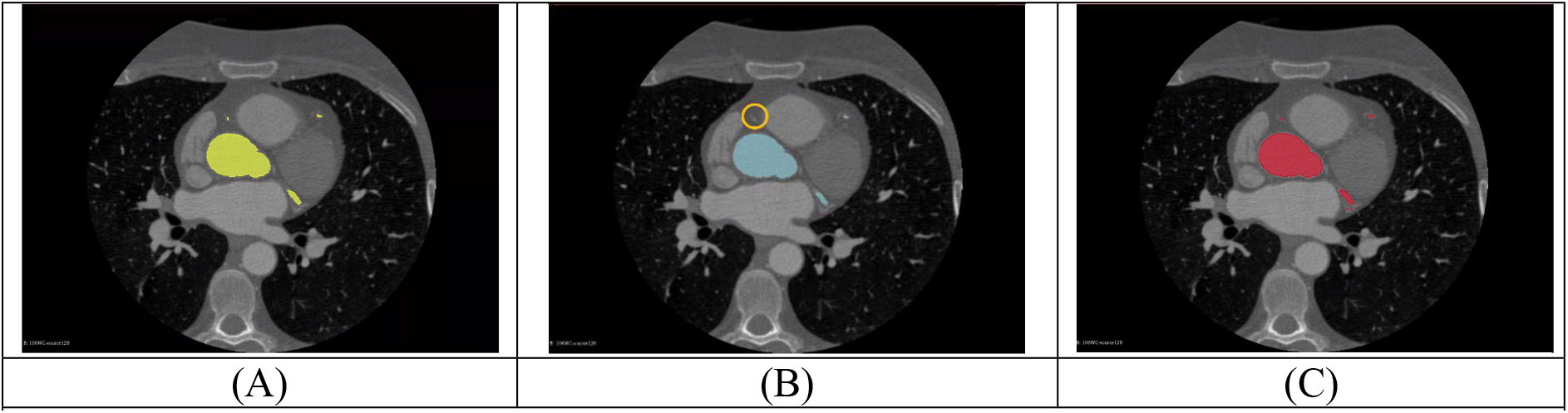
(A) Ground-truth mask (yellow) (B) Mask from Simpleware-ScanIP (blue) with a missing vessel (orange circle) (C) Mask from our model (red)

The segmentation results of patients 1 and 2 are displayed in Figures 7 and 8. From Figure 7, it is clear that our method can segment the aorta and coronary arteries, with a result very close to the optimal manual label. The Simpleware–ScanIP segments the aorta with good accuracy while some segments of the coronary arteries are missing.

**Figure 7:**
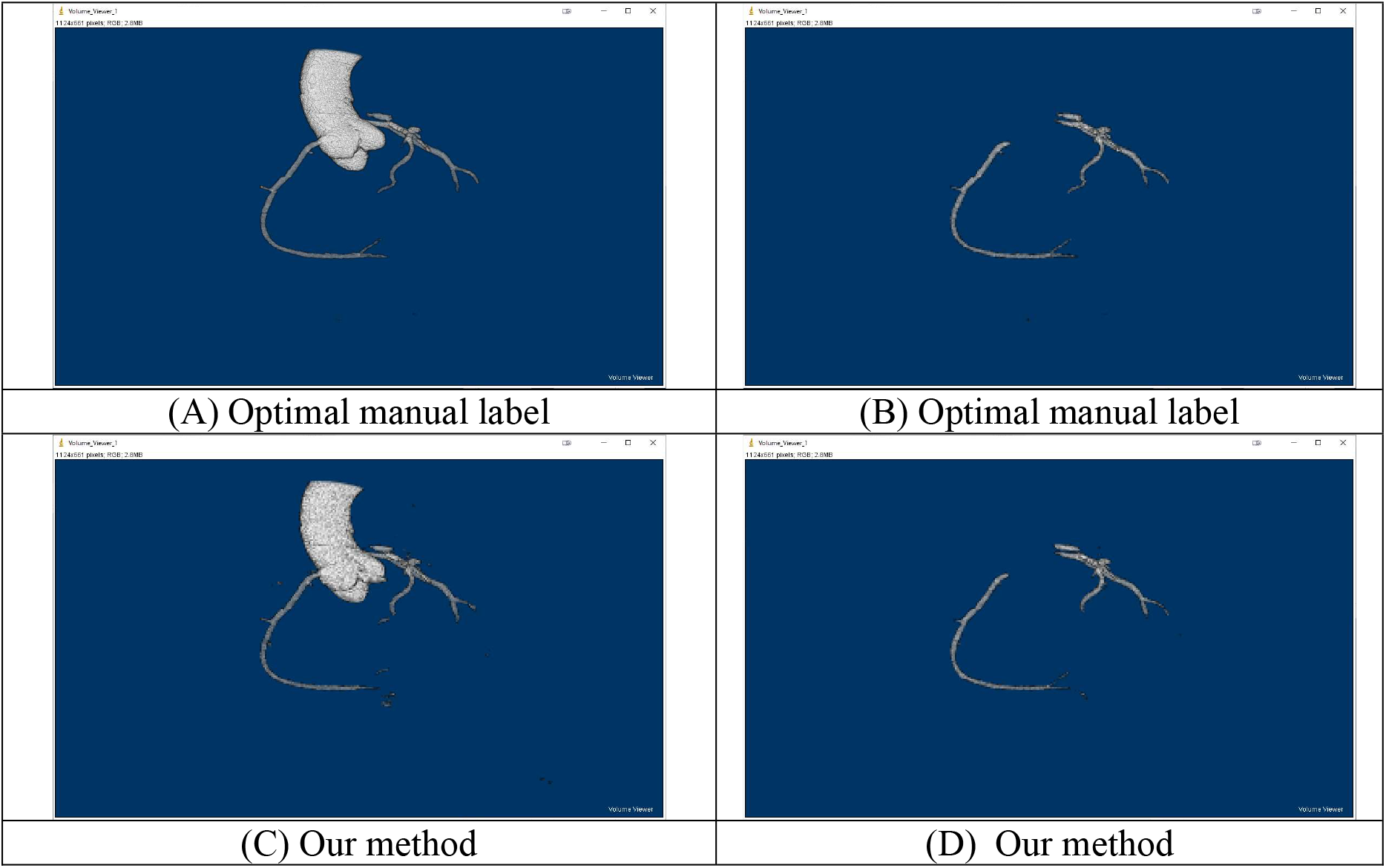
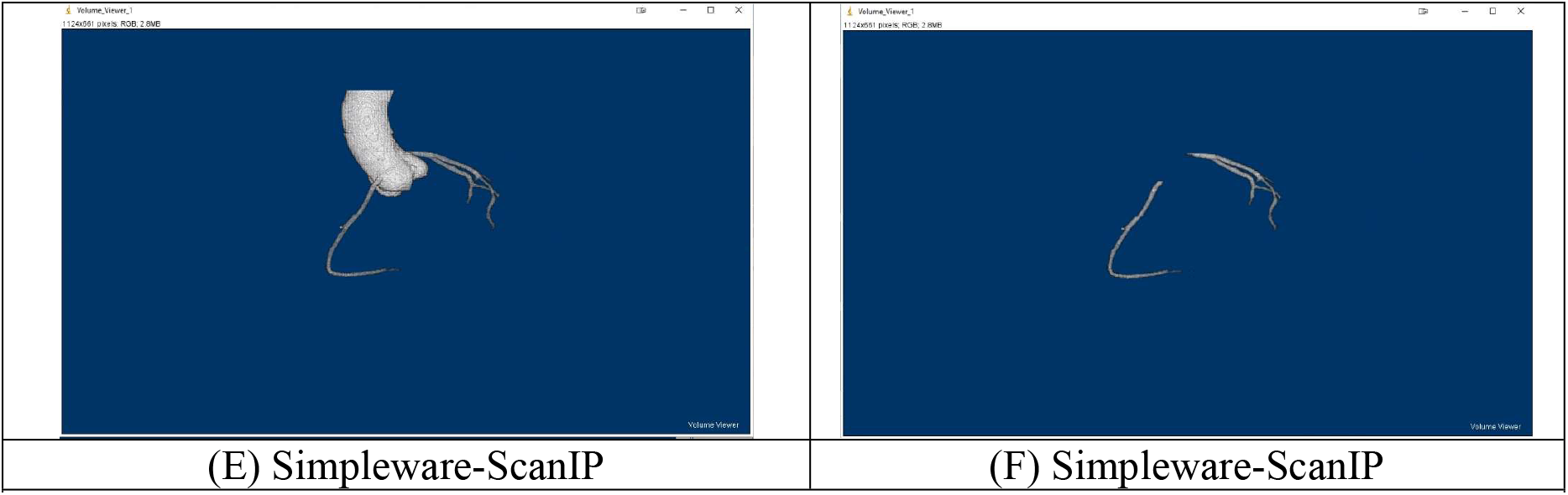
Segmentation results of Patient 1. Segmentation of aorta and coronary arteries: Optimal manual label, (C) Our method, (E) Simpleware–ScanIP. Segmentation of coronary arteries only: (B) Optimal manual label, (D) Our method, (F) Simpleware– ScanIP

**Figure 8:**
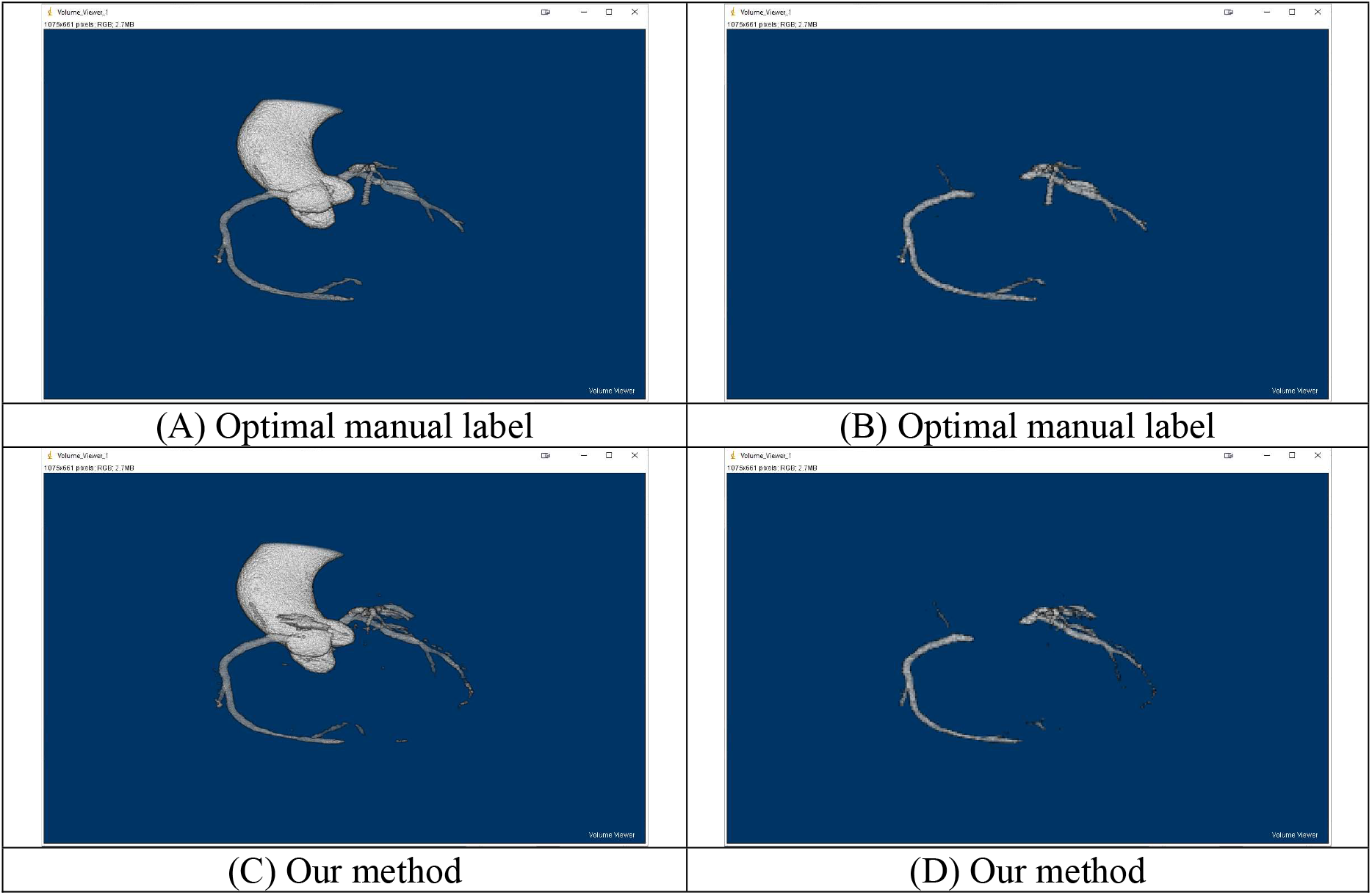
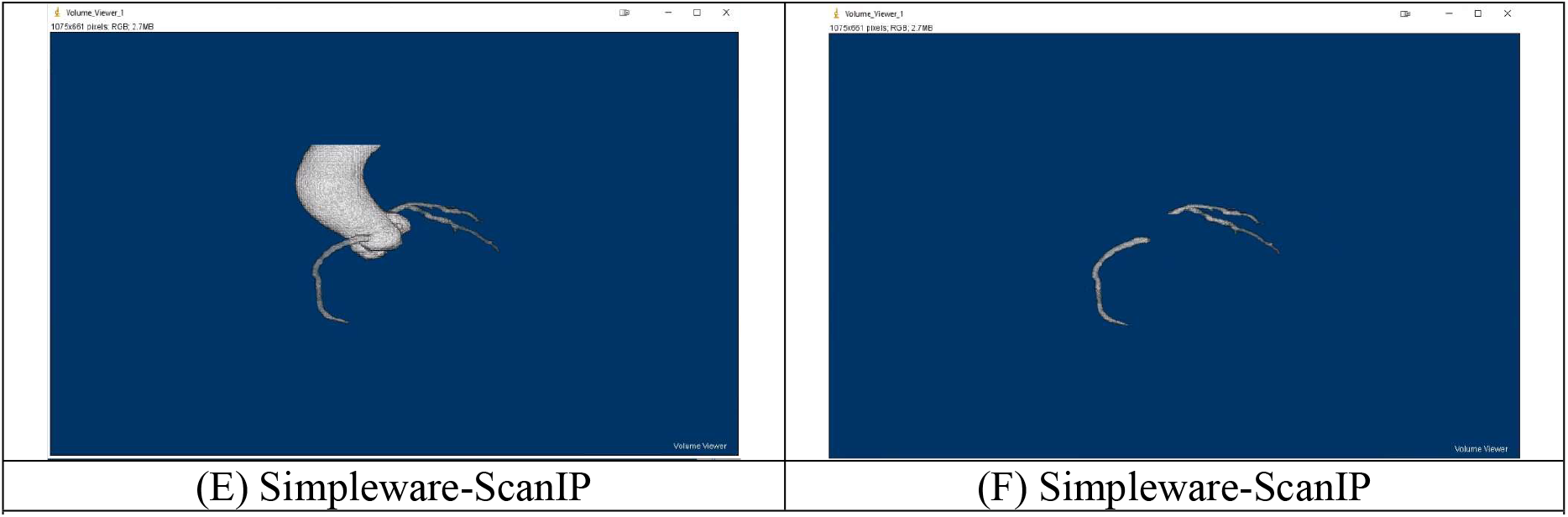
Segmentation results of Patient 2: with aorta and coronary arteries (A) Optimal manual label (C) Our method (E) Simpleware–ScanIP, with coronary arteries only (B) Optimal manual label (D) Our method (F) Simpleware–ScanIP

For patient 2 (Figure 8), our method can segment the aorta and coronary arteries well, but the segmentation incorporates an artefact. When segmenting the coronary arteries alone, some segments of the coronary arteries are missing in the segmentation. As expected, the aorta segmentation is good when using the semi-automatic method, while the segmentation of the coronary arteries is relatively poor. It should be noted that the artefact present in the segmentation of the aorta and coronary arteries using our method can be easily removed by excluding the non-connected components of the mask.

#### Segmentation time

The segmentation time of our proposed method and semi-automatic segmentation is shown in Table 3 (aorta and coronary arteries) and Table 4 (coronary arteries only) respectively. The mask prediction using our method was significantly faster than the Simpleware-ScanIP for both segmentation scenarios (*p*-value < 0.001), taking less than 4 seconds on average to predict the aorta and/or coronary arteries masks. Additionally, the segmentation time is between 40s and 141s when only a CPU with multi-cores is used.

**Table 3:** Segmentation time – mask contains aorta and coronary arteries

| Table 3: Segmentation time – mask contains aorta and coronary arteries |  |  |
| --- | --- | --- |
| With aorta | Time (average) | Time (SD) |
| Simpleware-ScanIP | 203.07s | 101.69s |
| Our method<br>(GPU) | 3.29s | 0.47s |
| Our method<br>(CPU only – 2 cores, 4 threads) | 138.57s | 15.74s |
| Our method<br>(CPU only – 16 cores, 32 threads) | 40.93s | 4.71s |

**Table 4:**
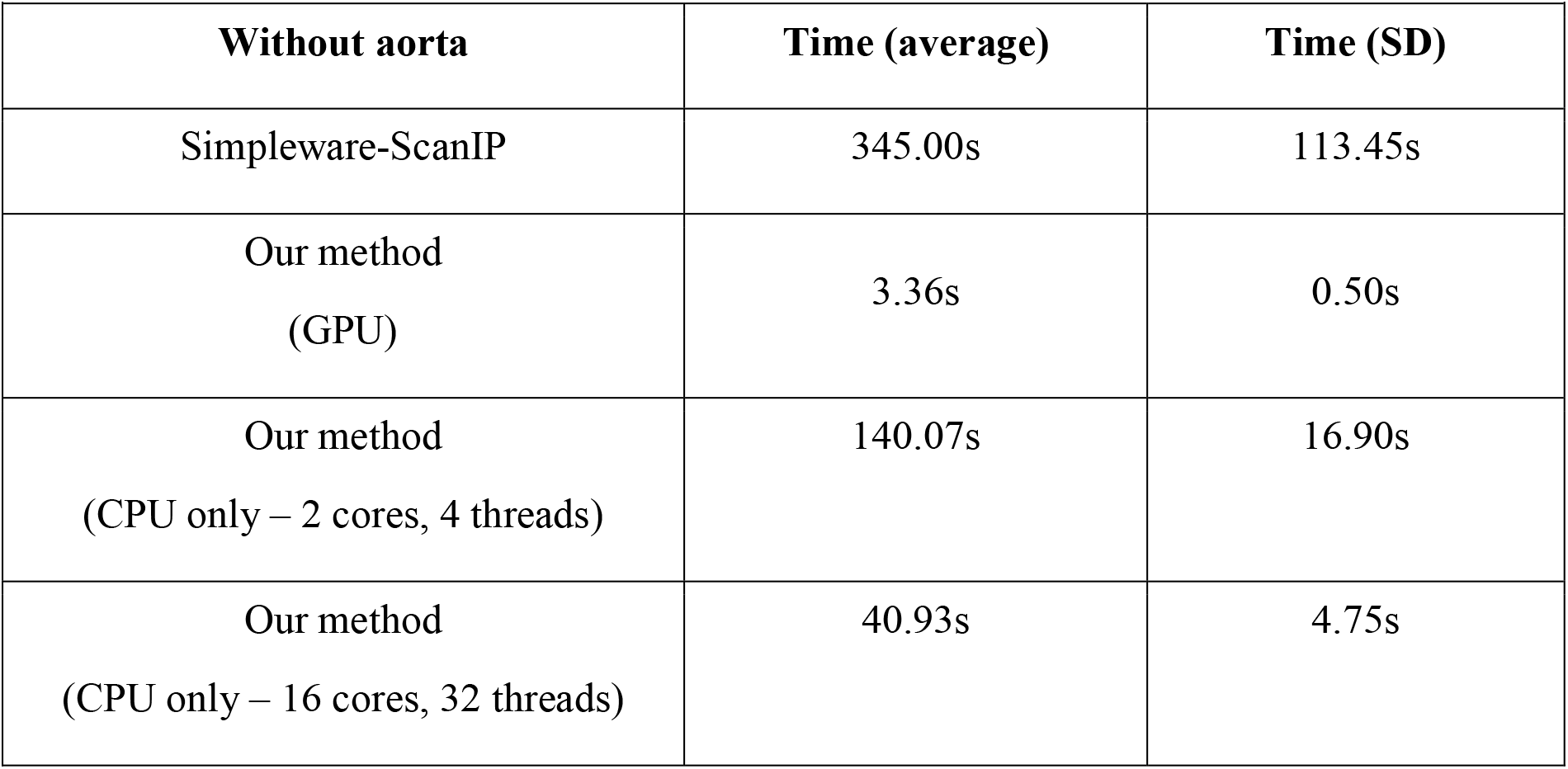
Segmentation time – mask contains coronary arteries only

## Discussion

A deep learning model based on a 2D Unet has been developed to segment the aorta and/or coronary arteries on cardiac CTCA images. Two models were trained to segment ROIs in two scenarios - (1) the aorta and coronary arteries (2) coronary arteries only. Our method demonstrates 91.20% and 88.80% DSC accuracy on scenarios 1 and 2 respectively. This suggests that our method can segment the aorta and/or coronary arteries with high accuracy.

Compared with a published 3D deep learning model [22] for scenario 1, which uses a 3D global feature embedded network with active contour loss, the performance of our method is similar (DSC: 91.20% (our method) vs 91.43% (ref. [22])). Our proposed method utilised a smaller number of network parameters resulting in more efficient training and a faster prediction time compared to the published 3D model. Our method requires less GPU memory, which is a common limitation when training and implementing a 3D model. It should also be noted that our method does not require GPUs for deployment, which favours its application in hospital networks where typically only CPUs are available.

The performance of our method was also compared with the 2D RCNN + 2D Unet technique [32] for scenario 2. Our method showed comparable performance (IoU: 79.85% (our method) vs 84.36% (ref. [32])), but does not require implementation of a sequence model to detect the coronary arteries. Implementing an additional sequence model would increase the computational complexity and hence decrease the algorithmic efficiency. It should be noted that the difference in reported accuracy between these methods may relate to the different test sets that were evaluated.

Compared with semi-automatic methods, our model performance is degraded when segmenting the aorta and coronary arteries. However, our model gives improved accuracy when segmenting the coronary arteries alone. The findings highlight the importance of evaluating segmentation performance of large vessels and small vessels separately to reduce the potential bias of segmentation performance metrics. In terms of the prediction time, our proposed model provided the fastest prediction when compared with the semi-automatic method. Though the time difference is statistically significant, the impact in time may be negligible from clinical perspective.

One advantage of our study is that a larger sample size (compared with published 2D approach) is used for training and prediction. This allows a more generalizable model can be obtained and therefore a more reliable prediction can be performed.

There are several limitations of this study. The design of the study is retrospective, and accordingly may suffer from patient selection bias. The ground-truth labels of this study were obtained by manual annotation and it is possible that the accuracy of the labels were potentially biased to the annotator’s experience. The performance of our model was compared with existing models using different datasets and we were not able to directly compare the various models on the same imaging dataset. Although our method can predict the segmented mask with good accuracy, visual inspection of the imaging by experts is still required. Currently, some regions of the proximal coronary artery are missed when using our models. Further improvements could be made by incorporating an attention gate to our model, which could allow the network to focus more closely on the coronary arteries during training.

## Conclusion

Our study demonstrates that a 2D UNET model is able to segment the coronary arteries efficiently and with good accuracy. It has the advantage that it can be deployed within hospital computer networks where GPUs are not available. Our study is a first essential stage of work to develop fully automatic detection and classification systems for CAD by using computer-based deep learning algorithms.

## Data Availability

Due to the nature of this research, participants of this study did not agree for their data to be shared publicly, so supporting data is not available.

## Acknowledgements

The authors would like to thank Mr Xiaoxia Yang for his assistance of data processing.

## References

[1] “World Health Organization, The Top 10 Causes of Death 2016 (https://www.who.int/news-room/fact-sheets/detail/the-top-10-causes-of-death)

[2] M. J. Bom, D. J. van der Heijden, E. Kedhi, J. van der Heyden, M. Meuwissen, P. Knaapen, et al., “Early Detection and Treatment of the Vulnerable Coronary Plaque: Can We Prevent Acute Coronary Syndromes?,” Circulation-Cardiovascular Imaging, vol. 10, May 2017.

[3] R. Senior, M. Monaghan, H. Becher, J. Mayet, and P. Nihoyannopoulos, “Stress echocardiography for the diagnosis and risk stratification of patients with suspected or known coronary artery disease: a critical appraisal. Supported by the British Society of Echocardiography,” Heart, vol. 91, pp. 427–436, Apr 2005.

[4] M. Saeed, T. A. Van, R. Krug, S. W. Hetts, and M. W. Wilson, “Cardiac MR imaging: current status and future direction,” Cardiovascular Diagnosis and Therapy, vol. 5, pp. 290–310, Aug 2015.

[5] N. Meyersohn, S. Janjua, P. Staziaki, D. Bittner, R. Takx, R. Weiner, et al., “Medical Management of Non-Obstructive Coronary Artery Disease on Coronary Ct Angiography: Insight from a Modern Emergency Department Clinical Registry,” Journal of the American College of Cardiology, vol. 67, pp. 1763–1763, Apr 5 2016.

[6] A. Wallis, N. Manghat, and M. Hamilton, “The role of coronary CT in the assessment and diagnosis of patients with chest pain,” Clinical Medicine, vol. 12, pp. 222–229, Jun 2012.

[7] “National Institute for Health and Care Excellence (NICE). Recent-onset chest pain of suspected cardiac origin: assessment and diagnosis Clinical guideline [CG95] https://www.nice.org.uk/guidance/cg95, 2016.

[8] F. Pugliese, M. G. M. Hunink, K. Gruszczynska, F. Alberghina, R. Malago, N. van Pelt, et al., “Learning Curve for Coronary CT Angiography: What Constitutes Sufficient Training?,” Radiology, vol. 251, pp. 359–368, May 2009.

[9] S. C. Saur, H. Alkadhi, P. Stolzmann, S. Baumuller, S. Leschka, H. Scheffel, et al., “Effect of reader experience on variability, evaluation time and accuracy of coronary plaque detection with computed tomography coronary angiography,” European Radiology, vol. 20, pp. 1599–1606, Jul 2010.

[10] P. D. Morris, A. Narracott, H. von Tengg-Kobligk, D. A. S. Soto, S. Hsiao, A. Lungu, et al., “Computational fluid dynamics modelling in cardiovascular medicine,” Heart, vol. 102, pp. 18–28, Jan 2016.

[11] W. F. Fearon, S. Achenbach, T. Engstrom, A. Assali, R. Shlofmitz, A. Jeremias, et al., “Accuracy of Fractional Flow Reserve Derived From Coronary Angiography,” Circulation, vol. 139, pp. 477–484, Jan 22 2019.

[12] Z. H. Li, J. Y. Zhang, L. Xu, W. J. Yang, G. Y. Li, D. X. Ding, et al., “Diagnostic Accuracy of a Fast Computational Approach to Derive Fractional Flow Reserve From Coronary CT Angiography,” Jacc-Cardiovascular Imaging, vol. 13, pp. 172–175, Jan 2020.

[13] L. Itu, S. Rapaka, T. Passerini, B. Georgescu, C. Schwemmer, M. Schoebinger, et al., “A machine-learning approach for computation of fractional flow reserve from coronary computed tomography,” Journal of Applied Physiology, vol. 121, pp. 42–52, Jul 1 2016.

[14] D. Bluestein, “Utilizing Computational Fluid Dynamics in Cardiovascular Engineering and Medicine-What You Need to Know. Its Translation to the Clinic/Bedside,” Artificial Organs, vol. 41, pp. 117–121, Feb 2017.

[15] C. Zhou, H. P. Chan, A. Chughtai, S. Patel, L. M. Hadjiiski, J. Wei, et al., “Automated coronary artery tree extraction in coronary CT angiography using a multiscale enhancement and dynamic balloon tracking (MSCAR-DBT) method,” Computerized Medical Imaging and Graphics, vol. 36, pp. 1–10, Jan 2012.

[16] R. Shahzad, H. Kirisli, C. Metz, H. Tang, M. Schaap, L. van Vliet, et al., “Automatic segmentation, detection and quantification of coronary artery stenoses on CTA,” International Journal of Cardiovascular Imaging, vol. 29, pp. 1847–1859, Dec 2013.

[17] I. Öksüz, D. Ünay, and K. Kadipasaoglu, “A Hybrid Method for Coronary Artery Stenoses Detection and Quantification in CTA Images,” 2012.

[18] Y. Kitamura, Y. Z. Li, W. Ito, and H. Ishikawa, “Coronary Lumen and Plaque Segmentation from CTA Using Higher-Order Shape Prior,” Medical Image Computing and Computer-Assisted Intervention - Miccai 2014, Pt I, vol. 8673, pp. 339-+, 2014.

[19] Y. Tian, Y. T. Pan, F. Q. Duan, S. F. Zhao, Q. J. Wang, and W. Wang, “Automated Segmentation of Coronary Arteries Based on Statistical Region Growing and Heuristic Decision Method,” Biomed Research International, vol. 2016, 2016.

[20] D. Lesage, E. D. Angelini, I. Bloch, and G. Funka-Lea, “A review of 3D vessel lumen segmentation techniques: Models, features and extraction schemes,” Medical Image Analysis, vol. 13, pp. 819–845, Dec 2009.

[21] “Clinical radiology UK workforce census report 2018 (https://www.rcr.ac.uk/publication/clinical-radiology-uk-workforce-census-report-2018)

[22] J. Gu, Z. Fang, Y. Gao, and F. Tian, “Segmentation of coronary arteries images using global feature embedded network with active contour loss,” Computerized Medical Imaging and Graphics, vol. 86, p. 101799, 2020/12/01/ 2020.

[23] W. Huang, L. Huang, Z. Lin, S. Huang, Y. Chi, J. Zhou, et al., “Coronary Artery Segmentation by Deep Learning Neural Networks on Computed Tomographic Coronary Angiographic Images,” Annu Int Conf IEEE Eng Med Biol Soc, vol. 2018, pp. 608–611, Jul 2018.

[24] C. Chen, C. Qin, H. Q. Qiu, G. Tarroni, J. M. Duan, W. J. Bai, et al., “Deep Learning for Cardiac Image Segmentation: A Review,” Frontiers in Cardiovascular Medicine, vol. 7, Mar 5 2020.

[25] P. Moeskops, J. M. Wolterink, B. H. M. van der Velden, K. G. A. Gilhuijs, T. Leiner, M. A. Viergever, et al., “Deep Learning for Multi-task Medical Image Segmentation in Multiple Modalities,” Cham, 2016, pp. 478–486.

[26] J. Merkow, A. Marsden, D. Kriegman, and Z. Tu, “Dense Volume-to-Volume Vascular Boundary Detection,” Cham, 2016, pp. 371–379.

[27] Ø. Kjerland, “Segmentation of Coronary Arteries from CT-scans of the heart using Deep Learning,” 2017.

[28] Y.-C. Chen, Y.-C. Lin, C.-P. Wang, C.-Y. Lee, W.-J. Lee, T.-D. Wang, et al., “Coronary Artery Segmentation in Cardiac CT Angiography Using 3D Multi-Channel U-net,” in MIDL 2019, 2019.

[29] Y. Shen, Z. J. Fang, Y. B. Gao, N. X. Xiong, C. S. Zhong, and X. H. Tang, “Coronary Arteries Segmentation Based on 3D FCN With Attention Gate and Level Set Function,” Ieee Access, vol. 7, pp. 42826–42835, 2019.

[30] M. C. H. Lee, K. Petersen, N. Pawlowski, B. Glocker, and M. Schaap, “TeTrIS: Template Transformer Networks for Image Segmentation With Shape Priors,” Ieee Transactions on Medical Imaging, vol. 38, pp. 2596–2606, Nov 2019.

[31] J. M. Wolterink, T. Leiner, and I. Išgum, “Graph Convolutional Networks for Coronary Artery Segmentation in Cardiac CT Angiography,” Cham, 2019, pp. 62–69.

[32] P. Mirunalini, C. Aravindan, A. T. Nambi, S. Poorvaja, and V. P. Priya, “Segmentation of Coronary Arteries from CTA axial slices using Deep Learning techniques,” Proceedings of the 2019 Ieee Region 10 Conference (Tencon 2019): Technology, Knowledge, and Society, pp. 2074–2080, 2019.

[33] Y. Lei, B. Guo, Y. Fu, T. Wang, T. Liu, W. Curran, et al., Automated coronary artery segmentation in Coronary Computed Tomography Angiography (CCTA) using deep learning neural networks vol. 11318: SPIE, 2020.

[34] O. Ronneberger, P. Fischer, and T. Brox, “U-Net: Convolutional Networks for Biomedical Image Segmentation,” Medical Image Computing and Computer-Assisted Intervention, Pt Iii, vol. 9351, pp. 234–241, 2015.

[35] C. A. Schneider, W. S. Rasband, and K. W. Eliceiri, “NIH Image to ImageJ: 25 years of image analysis,” Nat Methods, vol. 9, pp. 671–5, Jul 2012.

[36] A. Fedorov, R. Beichel, J. Kalpathy-Cramer, J. Finet, J. C. Fillion-Robin, S. Pujol, et al., “3D Slicer as an image computing platform for the Quantitative Imaging Network,” Magnetic Resonance Imaging, vol. 30, pp. 1323–1341, Nov 2012.

